# Is vestibular function related to human hippocampal volume?

**DOI:** 10.1101/2023.02.03.23285379

**Authors:** Joyce Bosmans, Hanne Gommeren, Peter zu Eulenburg, Annick Gilles, Griet Mertens, Angelique Van Ombergen, Patrick Cras, Sebastiaan Engelborghs, Vincent Van Rompaey

**Affiliations:** Experimental Laboratory of Translational Neurosciences and Dento-Otolaryngology, Faculty of Medicine and Health Sciences, University of Antwerp, Belgium; University Department of Otorhinolaryngology-Head and Neck Surgery, Antwerp University Hospital, Edegem, Belgium; German Center for Vertigo and Balance Disorders, University Hospital, Ludwig-Maximilians-University Munich, Munich, Germany; Graduate School of Systemic Neurosciences, Munich, Germany; Institute for Neuroradiology, University Hospital, Ludwig-Maximilians-University Munich, Munich, Germany; Department of Education, Health & Social Work, University College Ghent, Ghent, Belgium; Department of Neurology, Antwerp University Hospital and Born-Bunge Institute, University of Antwerp, Antwerp, Belgium; Department of Neurology, Universitair Ziekenhuis Brussel and Center for Neurosciences, Vrije Universiteit Brussel, Brussels, Belgium; Department of Biomedical Sciences, University of Antwerp, Antwerp, Belgium

**Keywords:** Hippocampus, Bilateral vestibulopathy, Hearing loss, Alzheimer’s disease, Cognition, Dementia

## Abstract

**OBJECTIVES:** Recent studies implicate the effect of vestibular loss on cognitive decline, including hippocampal volume loss. As hippocampal atrophy is an important biomarker of Alzheimer’s disease, exploring vestibular dysfunction as a risk factor for dementia and its role in hippocampal atrophy is of interest. The main objective is to replicate previous literature on whole-brain and hippocampal volumes in a group with bilateral vestibulopathy (BV).

**DESIGN:** Hippocampal and whole-brain MRI volumes were compared in adults aged between 55 and 83 years: (1) to substantiate previous literature, participants with BV (n=16) were compared to healthy controls (n=19), (2) to correct for a potential confounding effect of concomitant hearing loss, participants with BV were compared to healthy controls matched on age, sex, and hearing status (n=16), (3) to additionally evaluate the isolated effect of hearing loss on brain structure, participants with sensorineural hearing loss (SNHL; n=15) were compared to healthy controls. Furthermore, (4) to delineate otolith influence on hippocampal volume in a population with preserved vestibular function (healthy controls and SNHL combined; n=34), the role of saccular function was investigated.

**RESULTS:** Whole-brain and targeted hippocampal approaches using volumetric and surface-based measures yielded no significant differences in either of three comparisons: (1) BV versus controls, (2) BV versus matched controls, and (3) SNHL versus controls. Binary support vector machines were unable to classify inner ear health status above chance level. (4) Otolith parameters were not associated with hippocampal volume in a population with preserved vestibular function.

**CONCLUSIONS:** No significant differences in whole-brain or hippocampal volume were found when comparing BV participants with healthy controls, nor did concomitant SNHL confound this relationship. Saccular parameters in subjects with preserved vestibular function were not associated with hippocampal volume changes.

**Key points:**

- Recent research suggests an association between vestibular function and cognition.
- Hippocampal atrophy is an important biomarker of Alzheimer’s disease.
- Bilateral vestibular loss did not modulate hippocampal or whole-brain volume.

## 1. Introduction

Bilateral vestibulopathy (BV) is a severe chronic vestibular disorder of the labyrinth or the eighth cranial nerve characterized by postural imbalance, unsteadiness of gait which worsens in darkness and/or on uneven ground, and oscillopsia during head movements. Symptoms are typically absent under static conditions (Strupp et al., 2017). Multiple possible etiologies for BV exist, including but not limited to ototoxicity, bilateral Menière’s disease, bilateral vestibular schwannoma, genetic, or infectious causes (Lucieer et al., 2016).

There is evolving evidence suggesting that vestibular loss is associated with cognitive impairment and may even contribute to the onset of Alzheimer’s disease (Bigelow & Agrawal, 2015; Bosmans et al., 2022; Bosmans et al., 2021; Harun et al., 2016; Previc, 2013; Semenov et al., 2016).

When zooming in on the anatomical level, structural brain changes have been reported in patients with vestibular loss over the past twenty years in cross-sectional manual segmentation studies, specifically at the level of the hippocampus (Brandt et al., 2005; Hüfner et al., 2007). The hippocampus is a seahorse-shaped structure necessary for memory processing (encoding, consolidation, and retrieval) (Manns et al., 2003; Scoville & Milner, 1957) and spatial memory function (McNaughton et al., 1996; O’Keefe & Dostrovsky, 1971). These cognitive functions have been identified to be impacted in BV patients (Bosmans et al., 2022; Brandt et al., 2005; Dobbels, Mertens, et al., 2019; Dobbels, Peetermans, et al., 2019). Previous studies have compared hippocampal volumes between subjects with and without bilateral vestibular loss. Brandt et al. (2005) observed a significant selective shrinkage of hippocampal volume by 16.9% in people with BV relative to controls. A study by Kremmyda et al. (2016) described a significant reduction in grey-matter mid-hippocampal and posterior parahippocampal volume in long-standing BV patients compared to healthy controls. On the other hand, Göttlich et al. (2016) identified no association between hippocampal grey matter volume and BV. However, they observed bilaterally smaller hippocampal CA3 volumes with larger objective vestibulopathy-related disability, as measured by the Clinical Vestibular Score (Göttlich et al., 2016; Helmchen et al., 2009). A study by Dordevic et al. (2021) also observed a lack of volumetric differences in whole-brain and medial temporal lobe regions (including the hippocampus and insula) when comparing patients with chronic mild uni- or bilateral vestibulopathy and healthy controls. These findings are supported by Schöne et al. (2022) who analysed structural changes in the brain and hippocampus in patients with peripheral vestibular dysfunction, including a subgroup of BV patients. They also observed no hippocampal volume loss compared to age- and sex-matched healthy controls. Overall, studies evaluating hippocampal volume in bilateral vestibular loss have yielded contradicting results, with some studies observing an association between a decrease in hippocampal volume and BV (Brandt et al., 2005; Kremmyda et al., 2016) while other multi-site studies do not (Dordevic et al., 2021; Göttlich et al., 2016; Schöne et al., 2022).

A study by Kamil et al. (2018) took a different approach and evaluated hippocampal volume in healthy older adults (≥ 60 years) from the Baltimore Longitudinal Study of Aging (BLSA). They observed that a larger cervical vestibular-evoked myogenic potential (cVEMP) amplitude was significantly associated with a larger mean hippocampal volume (*p* = .003). They proposed that lower cVEMP amplitude, implying reduced saccular function, is significantly associated with a lower mean volume of the hippocampus. Jacob et al. (2020) included healthy older adults (≥ 60 years) from the BLSA cohort. They investigated the relation between vestibular function (using cVEMP) and the volume of structures comprised of or connected to the vestibular cortex. They observed smaller volumes of the hippocampus and entorhinal cortex associated with reduced vestibular function. A review by Smith (2019) supports these findings, stating that reduced saccular function can be associated with poorer spatial memory, Alzheimer’s disease, and reduced hippocampal volume.

Due to the close anatomical relationship in the inner ear, there is a high risk of concomitant sensorineural hearing loss (SNHL) in patients with vestibular dysfunction and vice versa (Lucieer et al., 2016; Zingler et al., 2007). As concomitant hearing loss could exacerbate a potential effect of vestibular dysfunction on brain volume, the hippocampus being of main interest, hearing levels should be included in these analyses. Previously mentioned studies comparing hippocampal volumes between BV patients and healthy controls generally lack a detailed description of hearing performance and did not include hearing performance in their methodological approach to the topic.

As we expect that concomitant SNHL might confound results from previous studies which found a decrease in hippocampal volume in the BV population, we hypothesize to find hippocampal changes in individual comparisons between people with BV and controls (not matched for hearing loss) and between subjects with SNHL and controls (both with preserved vestibular function). We hypothesize that the isolated effect of BV (adjusted for hearing level) will not result in significant hippocampal volume differences when compared to controls. This study aims to substantiate literature on hippocampal and whole-brain volumetric differences when comparing BV participants with healthy controls, adjusting for hearing level. Furthermore, an additional aim of this study is to delineate otolith influence on hippocampal volume in a population with preserved vestibular function. Therefore, there are four objectives of this study: (1) to compare whole-brain and hippocampal volumes between a group with BV and healthy controls, without matching for hearing status, and (2) to evaluate the isolated effect of bilateral vestibular loss on whole-brain and hippocampal volume by comparing a group with BV with an age-, sex-, and hearing-matched control group, (3) to separately evaluate the effect of hearing loss on whole-brain and hippocampal volume by comparing a group with SNHL and preserved vestibular function with an age- and sex-matched control group, and (4) to evaluate saccular function in a population with preserved vestibular function and its relation to hippocampal volume. In addition to hippocampal and whole-brain analyses, we will also perform cortical thickness and sulcus depth analyses for objectives 1 to 3 as well as surface-based morphometry.

## 2. Materials and Methods

### 2.1. Participant Characteristics

All participants were recruited from the *GECkO* study (Gehoor, Evenwicht, Cognitie), an ongoing prospective longitudinal cohort study of the effect of hearing loss and vestibular decline on cognitive function in older adults (Bosmans et al., 2020). This protocol was approved by the ethical committee of the University Hospital of Antwerp, Belgium (EC number B300201938949) and all participants gave their written informed consent in accordance with the Declaration of Helsinki prior to participation. The study protocol builds upon the Clinical Trials protocol with identifier NCT04385225.

#### 2.1.1. BV population

The diagnosis of BV was made according to the Bárány Society criteria and was defined as (1) a bilaterally pathological horizontal angular VOR gain (<0.6) measured by the vHIT, and/or (2) reduced horizontal angular VOR gain (<0.1) upon sinusoidal stimulation on a rotatory chair (0.1 Hz, Vmax = 50°/sec), and/or (3) reduced caloric response (sum of bi-thermal (30°C/44°C) maximum peak SPV on each side <6°/sec) (Strupp et al., 2017).

#### 2.1.2. Subjects with sensorineural hearing loss and healthy controls

People with hearing levels worse than age- and sex-based audiologically normal ranges in their best hearing ear were categorized in a group with SNHL. People with hearing levels within these audiologically normal ranges in their best hearing ear were categorized as healthy controls. Preserved hearing ranges were based on the ISO7029 method for air-conducting pure-tone threshold audiometry (frequencies 0.5, 1, 2, 3, 4, 6, 8 kHz). All participants underwent vHIT to confirm normal vestibular function (bilateral horizontal VOR gain > 0.6).

For all people (BV, SNHL, and healthy controls) the following inclusion criteria were applied (1) age 55 – 84 years, (2) Dutch as native language, (3) right-handed as defined by the Edinburgh Handedness Inventory (Oldfield, 1971), and (4) preserved cognitive function. A neuropsychological exam including a Mini-Mental State Examination (MMSE) and Repeatable Battery for the Assessment of Neuropsychological Status for Hearing impaired individuals (RBANS-H) was performed in all participants (Claes et al., 2016; Folstein et al., 1975). Participants were considered having preserved cognitive function when scoring ≥ 24/30 on the MMSE as well as ≥ percentile 16 on the RBANS-H total score (Albert et al., 2011; Folstein et al., 1975). People with an implanted hearing aid device (e.g., cochlear implant or bone-anchored hearing aid) were excluded from this study.

For the analyses comparing BV participants with healthy controls, adjusted for hearing level, BV participants were matched based on age, sex, and best aided speech audiometry in noise.

### 2.2. MRI Volumetry

#### 2.2.1. Acquisition Protocol

All subjects were investigated in a clinical 3.0 T scanner (Siemens Magnetom Prisma, Erlangen equipped with a 32-channel receiver head coil, 39 subjects in total, being 11 with BV, 10 with SNHL, and 18 healthy controls; Siemens Magnetom Vida, Erlangen equipped with a 64-channel receiver head coil, 11 subjects in total, being 5 with BV, 5 with SNHL, and 1 healthy control). A high-resolution T1-weighted image (GRAPPA sequence, 256 slices, slice thickness = 0.75 mm, voxel size = 0.75 x 0.75 x 0.75 mm, TR = 2060 ms, TE = 2.17 ms) was obtained in sagittal orientation.

#### 2.2.2. MRI Data Processing

Neuroimaging data quality control was performed via MRIQC version 0.15.1 (Esteban et al., 2017). Structural images were pre-processed and automatically segmented by the Computational Anatomy Toolbox (CAT12 Version 1980) (Figure 1, Panel A) (Gaser et al., 2022), an extension within the framework of Statistical Parametric Mapping software (SPM12) in MATLAB. Atlas-based segmentation for regions-based morphometry included the entire hippocampus as well as the volume of its substructures (CA1, CA2, CA3, dentate gyrus, and subiculum) taken from the cytoarchitectonic representation in the Julich Brain atlas (Amunts et al., 2020). In addition, total intracranial volume (TIV) was estimated and used (together with age and scanner type) as a covariate for all the voxel- and region-based, but not for surface-based analyses (Hutton et al., 2009).

**Figure 1.**
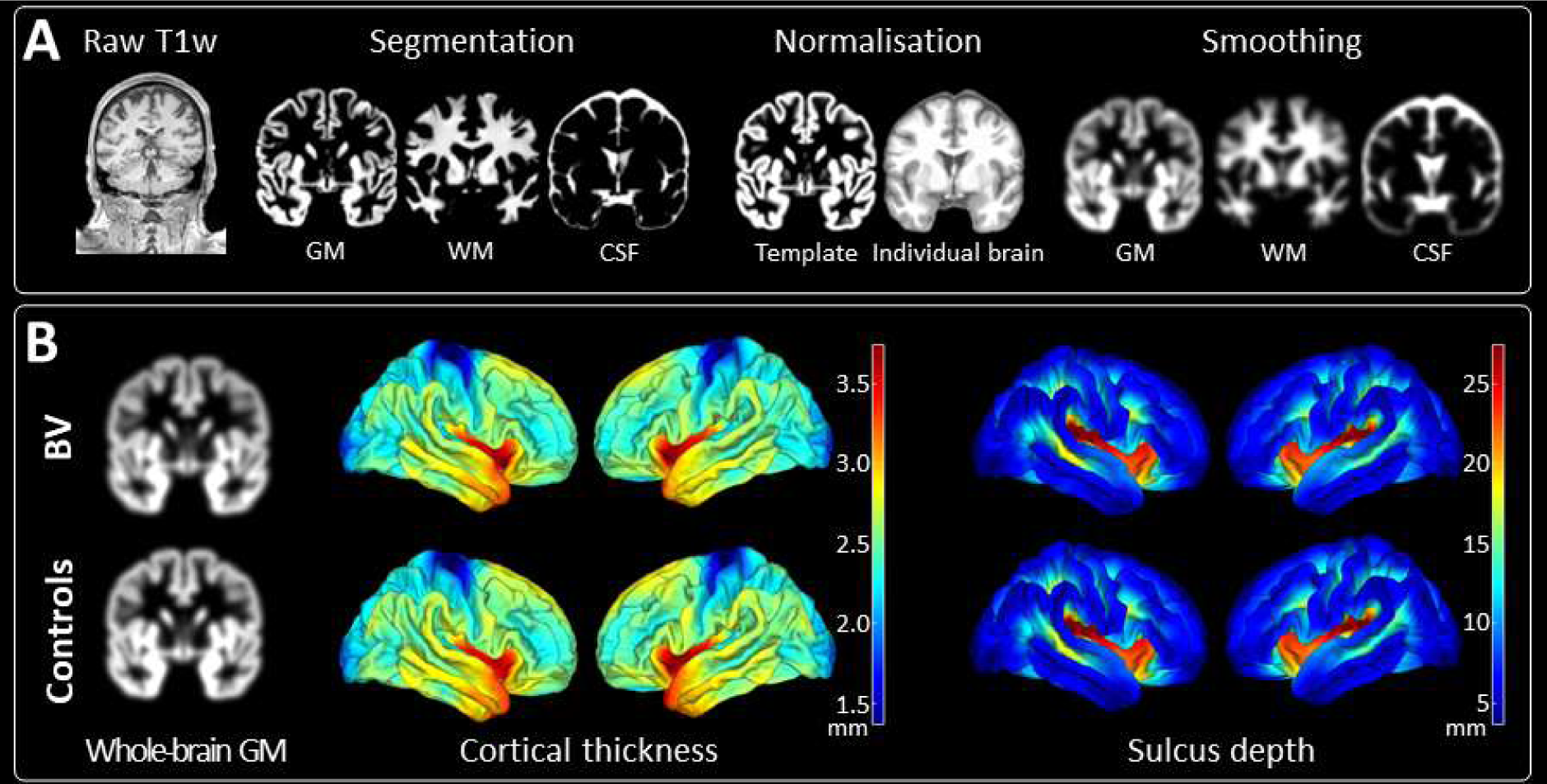
(A) Flowchart of the structural MRI preprocessing pipeline. All presented images are derived from the same control participant. The MNI152 NLIN 2009c 1mm template is used for normalisation. A smoothing kernel of 6mm full width at half maximum is applied. (B) Results of whole-brain comparisons between patients with BV (n=16) and their matched controls (n=16). Whole-brain comparisons encompassed whole-brain grey matter volumetric analyses and surface-based measures including cortical thickness and sulcus depth analyses. No significant differences were found in any of the comparisons. GM, grey matter; WM, white matter; CSF, cerebrospinal fluid; BV, bilateral vestibulopathy.

### 2.3. Otolith function evaluation of the saccule

Saccular function was investigated via the vestibulocollic reflex (VCR) using cVEMP with the validated Neuro-Audio device incorporating electromyography feedback (Neurosoft, DIFRA). While participants lay in a supine position, they lifted and rotated their head to one side, tensioning the sternocleidomastoid (SCM) muscle. Short 500 Hz tone bursts were presented in the contralateral ear at suprathreshold level (95 dB nHL). Present responses were biphasic and had two distinctive peaks (p13 and n23). Normative ranges were applied, with the p13 occurring 11.81–15.59 ms after stimulus onset, and with the n23 occurring 18.15–25.64 ms after stimulus onset (Li et al., 2014). Intact responses needed to be elicited at least twice to confirm presence of the VCR. Outcome measures included presence of intact responses (0, 1 ear, or both ears), and for each present response outcome measures included p13 latency (ms), n23 latency (ms), P-N amplitude (µV), rectified amplitude (µV), and SCM muscle contraction level (mean rectified voltage, MRV, µV).

### 2.4. Hearing Assessment

Unaided pure-tone audiometry was measured over a frequency range from 125 Hz to 8 kHz (specifically 0.125, 0.25, 0.5, 1, 2, 3, 4, 6, 8 kHz). Hearing thresholds were measured separately for each ear using a 2-channel Interacoustics AC-40 audiometer with insert earphones. Speech audiometry in noise (speech-in-noise; SPIN) was evaluated by the Leuven Intelligibility Sentences Test (LIST) with an adaptive procedure (van Wieringen & Wouters, 2008) in free field using a loudspeaker at a distance of 1 meter at 0° azimuth. The noise level was constant at 65 dB sound pressure level (SPL) while the speech level was adapted according to a correct (decreased speech level of 2 dB SPL) or incorrect (increased speech level of 2 dB SPL) response. Two lists of ten sentences each were conducted to acquire the speech reception threshold (SRT in dB SNR; averaged speech levels of the last five sentences and the imaginary 11^th^ sentence), both in an unaided and aided condition. The mean value of the best aided condition was used for analyses.

### 2.5. Statistical Analysis

For demographic and region of interest (ROI) based analyses (by use of the Julich-Brain atlas (Amunts et al., 2005)), JMP Pro 15 (Medmenham, UK) was used. Levene’s tests and visualization of data using histograms confirmed equal variances and the normality of reported data. However, because of the small sample size, nonparametric tests with the median and range are reported. Continuous patient characteristics were compared using Kruskal-Wallis ANOVA, for nominal patient characteristics, the Pearson Chi-squared statistic was used. For voxel-based morphometry analyses, the CAT12 toolbox and SPM12 were used. For each aim, a two-sample t-test was performed. Whole-brain changes were investigated by an F-contrast, with age, TIV, and scanner type as covariates. Similar statistics were performed for surface analyses (cortical thickness and sulcus depth), with only age and scanner type as covariates. Regarding *p*-value adjustment, the Monte-Carlo method for permutation testing (10.000 permutations) was applied using the TFCE toolbox (Version 224), with correction for multiple comparisons via false discovery rate (*p* < .05). In addition, machine learning in the form of multi-voxel pattern analysis is performed to increase the sensitivity to detect differences in each pairwise comparison by use of the Pattern Recognition for Neuroimaging Toolbox v3.0 (PRoNTo) (Schrouff et al., 2016). Classification was performed using a binary support vector machine (SVM) with one subject per class left out as the cross-validation scheme and 10.000 permutations. A Spearman correlation (and its 95% confidence interval) was performed for saccular analyses. *P*-values are reported, as well as *eta squared* (η²) indicating the effect size. The Pearson Chi-squared statistic was used for ordinal parameters, with *w* indicating its effect size. Between-scanner type differences were examined by a two-sample t-test of quality control parameters derived from MRIQC.

## 3. Results

### 3.1. Patient Characteristics

Demographic and clinical details as well as neuroimaging data quality of included participants can be found in Table 1 (BV versus healthy controls, without accounting for hearing loss and SNHL versus healthy controls, with preserved vestibular function) and Table 2 (BV versus age-, sex-, and hearing-matched controls). Sixteen patients with BV (median age = 63, range [56, 74], 10 males); 15 patients with SNHL (median age = 71, range [58, 83], 8 males); and 19 healthy controls (median age = 70, range [55, 81], 11 males) participated in the study. The median [range] disease duration for the BV population was 8 years [2, 22]. Among the etiologies of BV, 6 patients had a genetic risk (DFNA9), 1 patient autoimmune, 2 patients infectious (meningitis, varicella zoster), 1 patient ototoxic, 2 patients due to trauma, 1 patient with unknown etiology, and 3 patients idiopathic. All patients with idiopathic etiology had undergone an MRI internal auditory canal, tonal audiometry, and (hetero)anamnesis to exclude other causes. To confirm the diagnosis of BV, patients must meet at least one out of three of the Bárány Society criteria (Strupp et al., 2017). All three criteria (bilaterally reduced vHIT response, rotatory chair, and caloric testing) were met by 25% (n = 4) of people with vestibular loss. In 37.5% (n = 6), two out of three criteria were fulfilled, and the remaining 37.5% (n = 6) of people met one criterion. Based on the unaided tonal audiometry of the best hearing ear, 6 subjects with BV demonstrated age-normal hearing function, 4 had moderate SNHL, and 6 had severe SNHL.

**Table 1.**
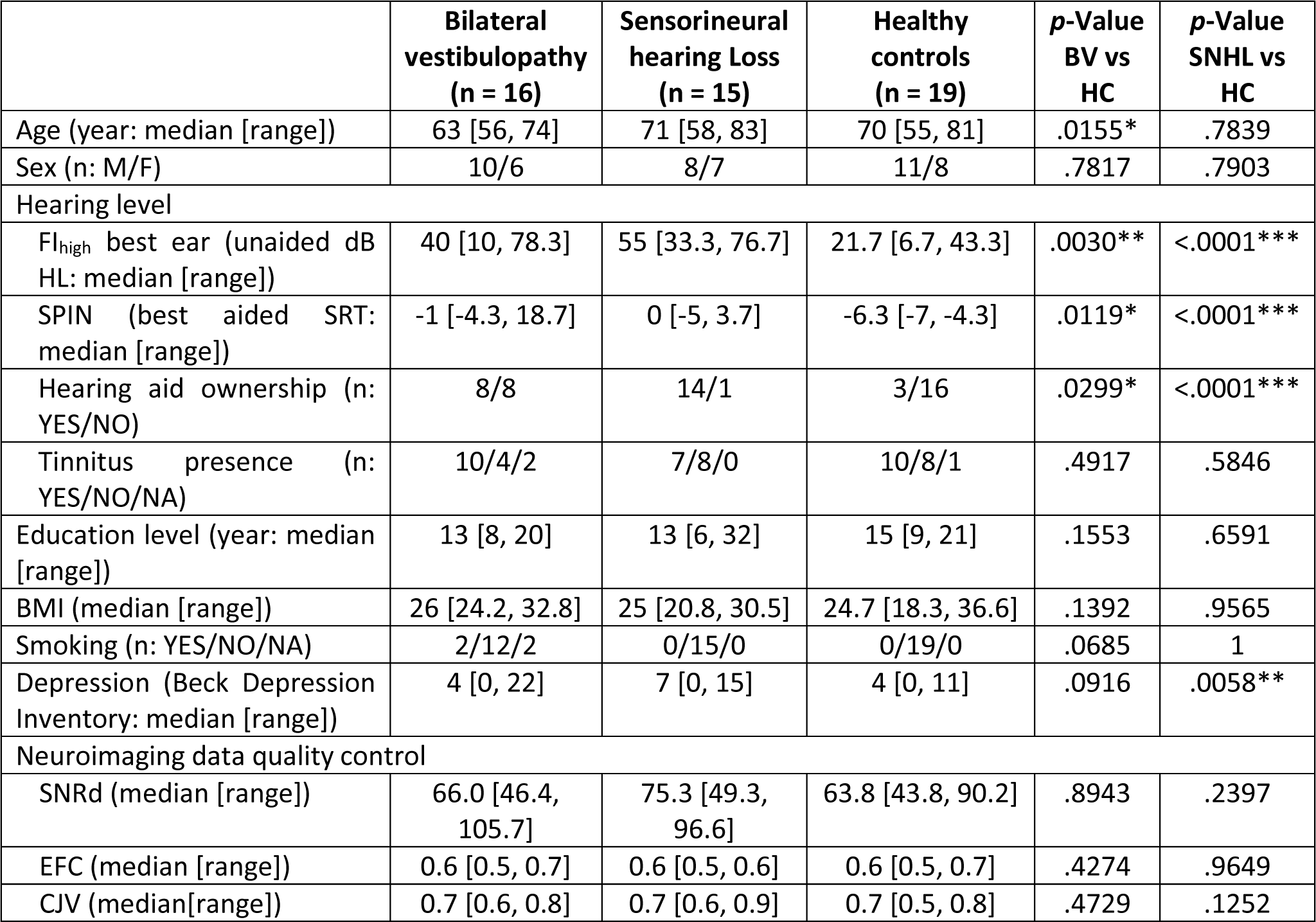
Demographic characteristics of people with BV, hearing loss, and healthy controls. Significant values are indicated with an asterisk (*: *p*<.05, **: *p*<.01, ***: *p*<.001). Education level indicates the years spent in school, starting from 6 years old. NA indicates the amount of missing data. BV, bilateral vestibulopathy; HC, healthy controls; SNHL, sensorineural hearing loss; SD, standard deviation; FI_high_, Fletcher index high (mean 1 – 2 – 4 kHz); dB HL, decibel hearing level; SPIN, speech-in-noise; SRT, speech reception threshold; BMI, body mass index; SNRd, Dietrich’s signal-to-noise ratio; EFC, entropy focus criterion; CJV, coefficient of joint variation.

**Table 2.**
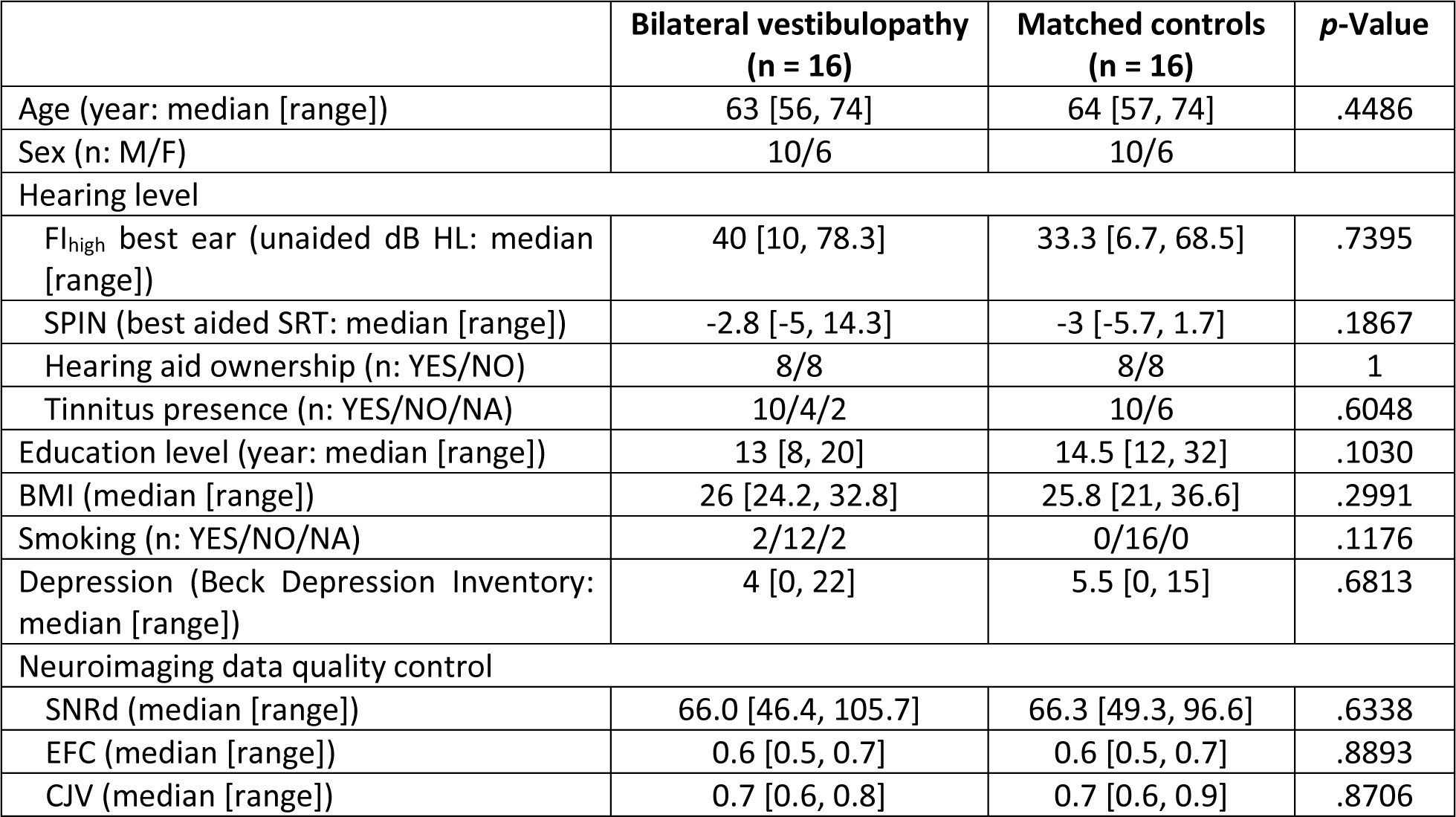
Demographic characteristics of people with BV and its age-, sex-, and hearing-matched controls. Education level indicates the number of years spent in school, starting from 6 years old. NA indicates the amount of missing data. SD, standard deviation; FI_high_, Fletcher index high (mean 1 – 2 – 4 kHz); dB HL, decibel hearing level; SPIN, speech-in-noise; SRT, speech reception threshold; BMI, body mass index; SNRd, Dietrich’s signal-to-noise ratio; EFC, entropy focus criterion; CJV, coefficient of joint variation.

Age, sex, hearing level, education level, obesity, smoking status, tinnitus presence, and depression may affect hippocampal volumes (Campbell et al., 2004; Cherbuin et al., 2015; Nobis et al., 2019; Profant et al., 2020). Therefore, age, sex, Fletcher index high (FIhigh; average threshold of 1 kHz, 2 kHz, and 4 kHz), SPIN, hearing aid ownership, years of education (number of years spent in school, starting from the age of 6 years old), body mass index (BMI), smoking status, tinnitus presence, and the total score of the Beck Depression Inventory were included in the demographic characteristics.

When comparing the BV group with healthy controls, not matched for hearing status, significant differences were observed for hearing level (Fletcher index high, SPIN, and hearing aid ownership), highlighting the importance of including hearing status in future analyses. In addition, significant differences were found for age (Table 1). These differences must be considered when interpreting results. When comparing the BV group with age-, sex-, and hearing-matched controls to single out the sole effect of bilateral vestibular loss, no significant demographic or patient characteristic differences were observed (Table 2). When comparing a group with SNHL with healthy controls, both with preserved vestibular function, expected significant differences were observed for hearing level (Fletcher index high, SPIN, and hearing aid ownership). In addition, these groups differed significantly on depressive status, which needs to be considered when interpreting results (Table 1).

Neuroimaging data quality control encompassed image quality metrics for structural images including Dietrich’s signal-to-noise ratio (SNRd) (Dietrich et al., 2007), entropy focus criterion (EFC) (Atkinson et al., 1997), and coefficient of joint variation (CJV) (Ganzetti et al., 2016). Neuroimaging data quality control was blinded for diagnostic categories and afterwards tested for group differences. The parameters EFC and CJV were included to control for the potential head motion differences between the groups during structural neuroimaging. None of the pairwise comparisons resulted in a significant difference on any of the image quality metrics (Table 1, Table 2). For inter-scanner differences, the same image quality metrics were compared between both scanners including all obtained data. The Siemens Magnetom Prisma was used in 39 participants with median [range] age 65 years [55, 81], including 21 males and 18 females, being 11 participants with BV, 10 with SNHL, and 18 healthy controls. The Siemens Magnetom Vida was used in 11 participants with median [range] age 72 years [63, 83], including 8 males and 3 females, being 5 participants with BV, 5 with SNHL, and 1 healthy control. There was no significant difference for SNRd and EFC between scanners (*p* = .7223; *p* = .8550; respectively). However, CJV differed significantly between scanners (*p* = .0009), with larger values and therefore more head motion in the Siemens Magnetom Vida scanner. This has to be kept in mind when interpreting results.

### 3.2. A comparison between the BV population and healthy controls, unaccounted for hearing status

To try and reproduce results from previous studies, tissue segmentation of people with BV was compared with healthy controls, unaccounted for hearing status. No suprathreshold clusters, thus no significant changes in whole-brain grey matter volume between these two groups were observed (*p* > .05). A dedicated ROI analysis of the hippocampus proper (CA1 – CA3) also revealed no significant differences between these two groups (*p* = .7137). Lateralization analyses of the hippocampus proper again found no significant difference between these two groups (left hippocampus proper: *p* = .6237; right hippocampus proper: *p* = .8562). Details of the ROI analysis of the hippocampus proper can be found in Table 3. Surface-based analyses (cortical thickness as well as sulcus depth) also found no significant difference between these two groups (*p* > .05). Machine learning is applied here as a more sensitive tool to detect differences between both groups. The area under the receiver operating characteristic (ROC) curve value of the SVM model remained 0 (*p* = 1) with a total accuracy of 55.26%, reflecting random classification of people with BV versus healthy controls unaccounted for hearing status, which is in line with previous results.

**Table 3.** ROI volumes of the hippocampus proper. BV, bilateral vestibulopathy; HC, healthy controls; SNHL, sensorineural hearing loss.

|  | Bilateral vestibulopathy | Sensorineural hearing loss | Healthy controls | Matched controls | p-Value BV vs HC | p-Value BV vs matched controls | p-Value SNHL vs HC |
| --- | --- | --- | --- | --- | --- | --- | --- |
| Left hippocampus proper (median [range]) | 3.3 [1.3, 3.8] | 2.9 [2.5, 3.8] | 3.1 [2.3, 3.6] | 3.2 [2.6, 3.8] | .6237 | .7200 | .7375 |
| Right hippocampus proper (median [range]) | 3.9 [3.1, 4.6] | 3.7 [2.9, 4.7] | 4.0 [2.9, 5.7] | 3.9 [3.4, 4.7] | .8562 | .8958 | .3271 |
| Hippocampus proper (median [range]) | 7.3 [5.1, 8.2] | 6.5 [5.4, 8.5] | 7.1 [5.4, 9.1] | 7.2 [6.0, 8.5] | .7137 | .7806 | .4520 |

### 3.3. Effect of bilateral vestibular loss on brain volumes

To evaluate the isolated effect of bilateral vestibular loss on brain tissue compartments and to exclude a potential confounding effect of concomitant hearing loss, modulated grey and white matter tissue volumes of people with BV were compared with healthy controls additionally matched for hearing status. Again whole-brain grey matter comparisons including substantial permutation testing to protect against false positive findings in small cohorts like ours yielded no significant differences between these two groups (*p* > .05) (Figure 1 Panel B). A ROI analysis of the hippocampus proper again found no significant morphometric changes between these two groups (total hippocampus proper: *p* = .7806; left hippocampus proper: *p* = .7200; right hippocampus proper: *p* = .8958; see Table 3; Figure 2). Surface-based analyses (cortical thickness and sulcus depth) also gave no significant differences between these two groups (*p* > .05) (Figure 1 Panel B). The SVM model resulted in an area under the ROC curve value of 0 (*p* = 1, total accuracy of 40.62%), reflecting again at random classification of people with BV versus their matched healthy controls, supporting these previous results.

**Figure 2.**
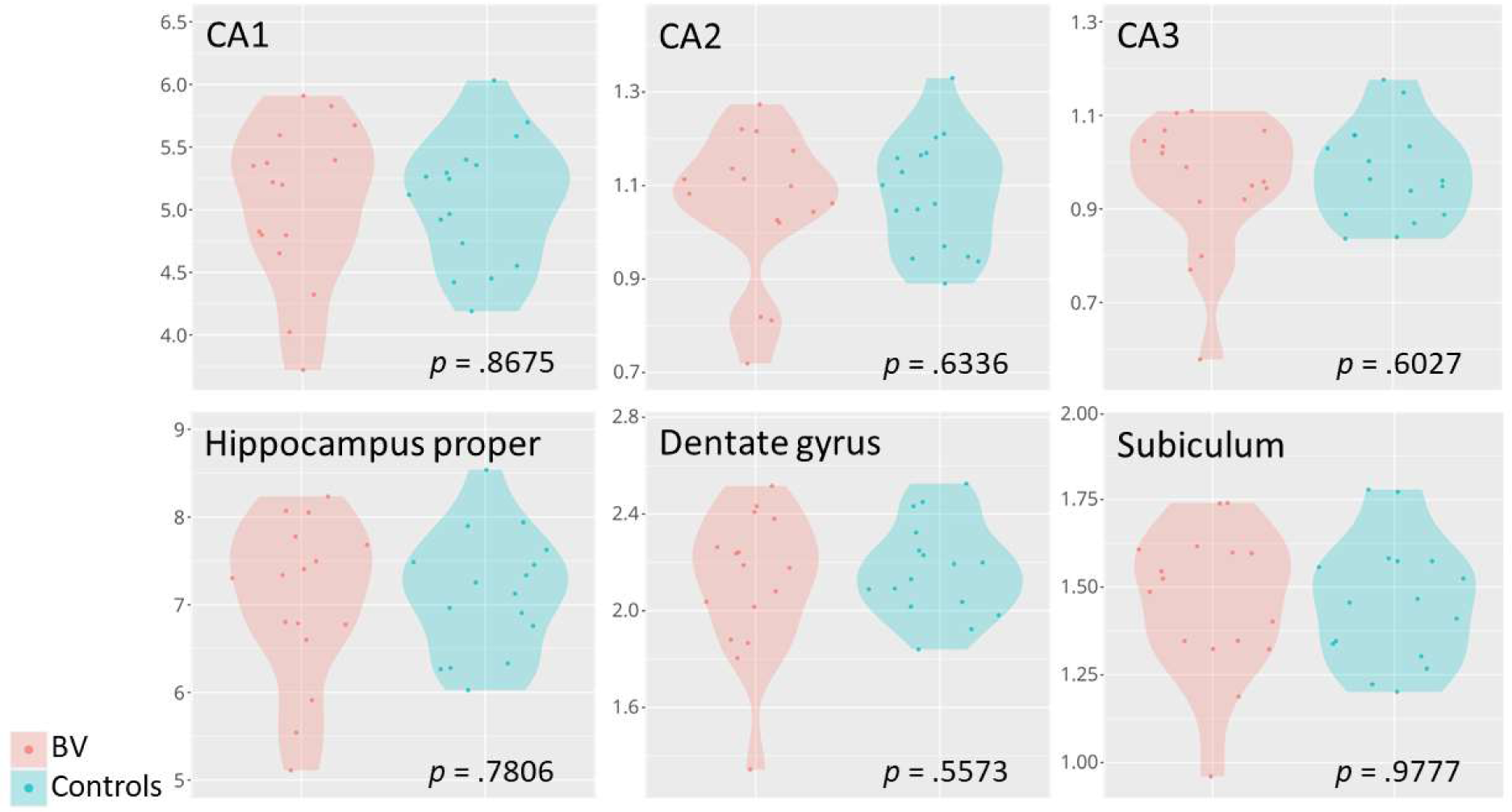
Targeted hippocampal volumetric measurements. Violin plots of the hippocampal subfields (in ml) of patients with BV (n=16) in comparison with their matched controls (n=16). The hippocampus proper is calculated as the sum of CA1, CA2, and CA3. BV, bilateral vestibulopathy; CA, cornu ammonis.

### 3.4. Effect of sensorineural hearing loss on brain volumes

To evaluate the sole effect of SNHL in voxel-based morphometry, in this case to explore the effect of this potential confounding factor when analyzing brain volumes of people with BV, brain volumes of subjects with SNHL were compared with healthy controls. Comparisons of whole-brain grey matter tissue found no significant differences between these two groups (*p* > .05), as well as a ROI analysis of the hippocampus proper (total hippocampus proper: *p* = .4520; left hippocampus proper: *p* = .7375; right hippocampus proper: *p* = .3271; see Table 3). In addition, surface-based analyses (cortical thickness and sulcus depth) yielded no significant differences between these two groups (*p* > .05). These results are in line with the SVM model which resulted in an area under the ROC curve value of 0 (*p* = 1, total accuracy of 65.79%).

### 3.5. Otolith (saccular) function and hippocampal volumes

To explore whether hippocampal volume correlates with saccular function in a population with preserved vestibular function, cVEMP parameters of participants without BV were analysed (Table 4). These analyses included a total of 34 participants (15 with SNHL and 19 healthy controls). Out of all 68 ears, 43 ears demonstrated an intact saccular response. However, the presence of intact responses was not significantly associated with the volume of the hippocampus proper (*X²*(2, *N* = 34) = .0804, *p* = .9606). Of the ears with intact responses, P-N amplitude, rectified amplitude, and n23 latency demonstrated no significant nor clinically meaningful effect (*r*(1) = −0.07, *p* = .643; *r*(1) = 0.01, *p* = .966; *r*(1) = 0.11, *p* = .472; respectively). Muscle tension of the SCM as measured by MRV also demonstrated no significant effect (*r*(1) = 0.16, *p*= .304). P13 latency on the other hand was significantly associated with hippocampal volume (*r*(1) = 0.34, *p* = .028) with a medium effect (η² = .1129). Even though cVEMP testing does not depend on hearing level but to correct for SNHL, p13 latency was correlated with unaided FIhigh-values of the best hearing ear (Rosengren et al., 2019). As expected, this correlation was not significant (*r*(1) = −0.001, *p* = .995) with a trivial effect size (η² < .001). There are heterogeneous results on the effect of age on p13 latency, but p13 latency is generally known to be associated with age (Macambira et al., 2017). Indeed, when including age and p13 latency as independent variables with total hippocampal volume as the dependent variable, this model was significant (F(2, 40) = 5.8485, *p* = .006). Parameter estimates were *p* = .020 for age and *p* = 0.107 for p13 latency. When removing p13 latency from this model, thus resulting in the correlation between total hippocampal volume and age, this model was significant (*r*(1) = −0.310, *p* = .010).

**Table 4.** Saccular characteristics and their association with volume of the hippocampus proper. Latencies are expressed in milliseconds, amplitude and muscle tension are expressed in microvolts. Significant results are indicated with an asterisk (*: *p*<.05). *p*-Values and effect sizes (uncorrected) are presented together with *p*-values and effect sizes corrected for age as a confounder. cVEMP, cervical vestibular-evoked myogenic potentials; MRV, mean rectified voltage.

| cVEMP parameter | Median [range] | Correlation with hippocampal volume (95% confidence interval) | *p*-Value uncorrected | *p*-Value corrected for age | Uncorrected effect size $\eta^2$ | Effect size $\eta^2$ corrected for age |
| --- | --- | --- | --- | --- | --- | --- |
| Presence of intact responses (n = 34) | No responses: n=6 (18%)<br>One ear: n=13 (38%)<br>Both ears: n=15 (44%) | Chi-Square (df=2):<br>.0804 | .9606 | .9382 | w = .0486 (trivial) | w = <.0001 (trivial) |
| P-N amplitude (n=43) | 102.5 [38.5, 195.2] | 0.07 (-0.23, 0.37) | .6429 | .8124 | .0053 (trivial) | .0012 (trivial) |
| Rectified amplitude (n=43) | 0.69 [0.36, 1.47] | 0.01 (-0.29, 0.31) | .9660 | .7502 | .00004 (trivial) | .0021 (trivial) |
| p13 latency (n=43) | 13.4 [12, 15.2] | 0.34 (0.04, 0.58) | .0276* | .1071 | .1129 (medium) | .0526 (small) |
| n23 latency (n=43) | 22 [18, 25.3] | 0.11 (-0.19, 0.40) | .4718 | .3754 | .0127 (small) | .0163 (small) |
| MRV (n=43) | 149.9 [90.5, 204.7] | 0.16 (-0.15, 0.44) | .3039 | .2173 | .0258 (small) | .0312 (small) |

## 4. Discussion

This study aimed to substantiate the literature on hippocampal and whole-brain volumetric differences when comparing BV participants with healthy controls whilst adjusting for hearing level. Previous studies on this inner ear topic did not control for the confounding effects of altered hearing levels. Hence, we first tried to reproduce previous literature by comparing a group of subjects with BV with a healthy control group, without considering hearing status. However, we were unable to find any structural differences between both groups: neither using whole-brain grey matter analyses, nor using an ROI analysis of the hippocampus proper, nor using surface-based analyses, nor using the SVM model as a more sensitive machine learning technique. As we hypothesized to observe a difference between these groups, we wanted to untangle the individual impact of vestibular dysfunction and potential concomitant hearing loss in this expected volumetric difference. The association between the isolated effect of BV and whole-brain and hippocampal volume resulted in no significant differences, which was in line with our hypothesis. However, we did expect to find a significant association when separately evaluating the effect of hearing loss on whole-brain and hippocampal volume, but this comparison also resulted in no significant difference.

In summary: in this study, neither BV nor SNHL was significantly associated with whole-brain or hippocampal volumetric differences in comparison with healthy controls. In addition, we aimed to delineate otolith influence on hippocampal volume in a population with preserved vestibular function. An intact cVEMP response was elicited in at least one ear in 82% of the cases. The p13 latency was positively correlated with hippocampal volume, where longer latencies within normal ranges indicated larger hippocampal volumes. However, when correcting for age, this significant correlation disappeared and could thus be explained by age as a confounding variable. Other saccular parameters at suprathreshold level (95 dB nHL) including the number of intact responses, P-N amplitude, rectified amplitude, n23 latency, and MRV did not demonstrate a significant correlation with the volume of the hippocampus proper.

This study used the normative ranges of Li et al. (2014) to indicate the presence of intact cVEMP responses (p13: 11.81-15.59 ms; n23: 18.15-25.64 ms). However, different latencies can be observed in the literature, with some diverging from the normative ranges of Li et al. (2014) (for a recent systematic review with meta-analysis, see Macambira et al. (2017)). For transparency reasons, an overview per subject of saccular parameters and additional relevant data can be found in Appendix A.

The emerging theory of the association between vestibular loss and cognitive decline would be supported by associated hippocampal atrophy in BV. As such, positive studies by Brandt et al. (2005) and Kremmyda et al. (2016) are often cited exclusively to substantiate this hypothesis. However, the role of the replication crisis should not be underestimated and these current null findings, together with those observed by Dordevic et al. (2021), Göttlich et al. (2016), and Schöne et al. (2022) need to be taken into account to correct earlier underpowered findings using less reliable segmentation approaches to avoid future false understandings of this association. However, one can question whether the present study’s negative results can completely disprove the association between hippocampal atrophy and BV? Not necessarily. First of all, BV is a broad and heterogeneous condition. Therefore, one might consider subdividing the BV population by etiology or duration since onset. Second, multiple tests exist to assess peripheral vestibular end-organ functioning. The current study included older adults diagnosed with BV. Diagnostic criteria for this condition all rely on semicircular canal function. However, measurements of otolithic organs may be of added value. They may provide interesting new insights because of their association with spatial learning and memory (Smith, 2019). Therefore, this study included saccular characteristics and their association with hippocampal volume. Even though no association between saccular function and brain volumetry was observed, a previous systematic review described longer p13 latencies and smaller VEMP amplitudes with increasing cognitive decline along the Alzheimer’s disease continuum (Bosmans et al., 2021). It appears that the association between vestibular dysfunction and an increased risk of Alzheimer’s disease remains on a behavioral level and is not expressed at the anatomical level.

One thing that must be kept in mind is the sample size. Our research included 16 participants with BV, 15 with SNHL, and 19 healthy controls. Although as a rule of thumb, it is recommended that each subgroup should include at least 20 participants (Gaus & Rainer, 2013). However, we believe that the obtained data quality and stringency of the employed processing pipeline together with the application of full permutation testing makes our findings extremely robust.

A minor limitation is the difference in disease duration for the current BV population. Our study’s median [range] disease duration was 8 [2-22] years. Comparable studies have a disease duration of 5-10 years (Brandt et al., 2005), 13.6 ± 17.4 years (Kremmyda et al., 2016), and 3 months to 20 years (Göttlich et al., 2016). The high variation in disease duration might complicate a direct comparison between studies.

Another limitation could be the influence of confounding factors within our analyses. When comparing BV with healthy controls not accounted for hearing status, a significant difference was observed in age (with healthy controls being older). This is important because two groups are ideally comparable in age. If the BV group ages a few years such that they are comparable to the healthy controls regarding age, the BV group might present with smaller hippocampal volumes, considering that hippocampal volume tends to decrease with age. Therefore, the difference in hippocampal volume between BV patients and healthy controls might be more pronounced than demonstrated in this manuscript. When comparing the SNHL group with healthy controls, a significant difference was observed in depression, with the SNHL group obtaining higher depression scores. These significant confounding factors need to be kept in mind when interpreting results.

## 5. Conclusion

Neither whole-brain nor hippocampal volume differences were observed when comparing subjects with BV and healthy controls. Saccular function testing in subjects with preserved semicircular canal function resulted in no significant correlations with hippocampal volume. The association between vestibular dysfunction and an increased risk of cognitive dysfunction may only be present on the behavioral level and may not be expressed at the anatomical level.

## Data Availability

The data that support the findings of this study are available from the corresponding author upon reasonable request.

## Acknowledgments

This work was supported by a Fonds voor Wetenschappelijk Onderzoek (FWO) Fundamental Research Project (Grant Number G042819N3).

## Data-availability statement

**Appendix A.** Overview per subject of sex, age, hearing level, saccular parameters, and hippocampal volume. All cVEMP latencies lying between the normative ranges of Li et al. (2014) and therefore included in the analyses are shaded in grey. NR indicates no response was found. FI_high_, Fletcher index high (mean 1 – 2 – 4 kHz, unaided, best hearing ear); cVEMP, cervical vestibular-evoked myogenic potential; MRV, mean rectified voltage; NR, no response.

| ID | Sex | Age | FI<sub>high</sub><br>best<br>ear | cVEMP right ear |  |  |  |  | cVEMP left ear |  |  |  |  | Hippocampal<br>volume |
| --- | --- | --- | --- | --- | --- | --- | --- | --- | --- | --- | --- | --- | --- | --- |
|  |  |  |  | P13<br>latency | N23<br>latency | P-N<br>amplitude | Rectified<br>amplitude | MRV | P13<br>latency | N23<br>latency | P-N<br>amplitude | Rectified<br>amplitude | MRV |  |
| 1 | Female | 76-80 | 43.33 | 12.3 | 19.6 | 165.0 | 1.10 | 149.8 | 13.4 | 18.0 | 66.1 | 0.43 | 153.4 | 5.35 |
| 2 | Female | 71-75 | 21.67 | 13.2 | 25.3 | 159.3 | 1.20 | 132.8 | 14.6 | 24.1 | 172.4 | 1.47 | 117.4 | 7.29 |
| 3 | Female | 61-65 | 33.33 | 14.3 | 22.0 | 62.1 | 0.51 | 121.6 | 15.0 | 21.9 | 80.7 | 0.57 | 142.5 | 6.76 |
| 4 | Female | 61-65 | 33.33 | NR | NR | NR | NR | NR | 14.0 | 24.1 | 107.1 | 0.69 | 155.0 | 6.26 |
| 5 | Male | 76-80 | 21.67 | 15.2 | 22.0 | 146.7 | 0.77 | 190.1 | 11.6 | 15.7 | 67.5 | 0.38 | 175.9 | 9.10 |
| 6 | Male | 66-70 | 30.00 | 14.7 | 24.7 | 101.6 | 0.77 | 131.9 | NR | NR | NR | NR | NR | 7.07 |
| 7 | Male | 61-65 | 31.67 | 16.0 | 25.2 | 178.1 | 1.12 | 158.5 | 14.2 | 24.0 | 85.7 | 0.53 | 162.7 | 7.13 |
| 8 | Male | 76-80 | 31.67 | 14.0 | 23.8 | 99.2 | 0.66 | 151.4 | 13.1 | 19.5 | 64.3 | 0.48 | 132.7 | 7.98 |
| 9 | Female | 71-75 | 28.33 | NR | NR | NR | NR | NR | 12.8 | 20.8 | 38.5 | 0.37 | 103.5 | 6.23 |
| 10 | Male | 56-60 | 6.67 | 14.0 | 23.0 | 130.5 | 0.88 | 148.0 | 14.4 | 19.7 | 78.1 | 0.43 | 180.5 | 7.25 |
| 11 | Female | 51-55 | 15.00 | 15.7 | 23.4 | 71.3 | 0.53 | 134.2 | 12.2 | 16.9 | 68.9 | 0.58 | 119.1 | 7.06 |
| 12 | Female | 56-60 | 21.67 | 14.8 | 20.7 | 62.0 | 0.56 | 111.3 | 13.4 | 21.0 | 100.5 | 0.77 | 131.3 | 7.49 |
| 13 | Female | 71-75 | 15.00 | 19.3 | 25.4 | 71.5 | 0.48 | 147.6 | 15.6 | 24.3 | 65.0 | 0.54 | 121.4 | 5.57 |
| 14 | Male | 66-70 | 25.00 | 11.2 | 18.6 | 108.4 | 0.69 | 156.5 | 14.0 | 23.4 | 59.8 | 0.61 | 147.9 | 7.90 |
| 15 | Male | 71-75 | 28.33 | NR | NR | NR | NR | NR | 13.2 | 21.6 | 69.4 | 0.77 | 90.5 | 5.83 |
| 16 | Male | 56-60 | 16.67 | 14.9 | 21.4 | 136.3 | 0.90 | 150.7 | 13.1 | 20.0 | 140.9 | 0.69 | 204.7 | 7.33 |
| 17 | Female | 66-70 | 20.00 | 12.3 | 23.4 | 154.8 | 0.99 | 156.6 | 13.1 | 22.6 | 184.3 | 1.02 | 180.7 | 6.34 |
| 18 | Female | 66-70 | 16.67 | 12.4 | 22.2 | 100.5 | 0.64 | 156.1 | 13.4 | 24.9 | 163.8 | 0.98 | 167.6 | 7.23 |
| 19 | Male | 81-85 | 26.67 | 12.3 | 20.5 | 102.5 | 0.58 | 176.5 | 12.7 | 19.7 | 151.8 | 0.84 | 180.9 | 5.92 |
| 20 | Male | 71-75 | 15.00 | 15.2 | 21.2 | 112.0 | 0.67 | 168.2 | 17.3 | 21.8 | 84.7 | 0.54 | 156.3 | 6.33 |
| 21 | Male | 61-65 | 18.33 | 12.6 | 22.2 | 106.2 | 0.61 | 173.2 | 12.0 | 20.5 | 128.0 | 0.81 | 157.8 | 7.45 |
| 22 | Male | 61-65 | 45.00 | 13.8 | 22.5 | 99.6 | 0.97 | 103.1 | 14.0 | 22.0 | 148.3 | 1.08 | 137.0 | 6.28 |
| 23 | Male | 81-85 | 46.67 | 12.7 | 22.9 | 141.0 | 0.96 | 147.2 | 13.1 | 23.2 | 97.8 | 0.59 | 164.4 | 5.60 |
| 24 | Male | 71-75 | 55.00 | 13.4 | 23.3 | 85.0 | 0.61 | 139.0 | 12.7 | 23.2 | 130.6 | 0.87 | 149.9 | 6.91 |
| 25 | Male | 56-60 | 53.33 | 13.2 | 20.4 | 104.6 | 0.70 | 149.9 | NR | NR | NR | NR | NR | 7.94 |
| 26 | Female | 76-80 | 53.33 | 12.7 | 23.0 | 112.9 | 0.76 | 149.2 | 23.0 | 30.2 | 81.3 | 0.57 | 141.8 | 5.40 |
| 27 | Female | 56-60 | 53.33 | 13.4 | 20.8 | 195.2 | 1.20 | 162.7 | 18.4 | 24.5 | 71.2 | 0.44 | 161.9 | 7.63 |
| 28 | Male | 71-75 | 65.00 | 14.8 | 25.3 | 70.5 | 0.51 | 138.0 | 26.2 | 35.6 | 101.4 | 0.63 | 160.8 | 6.41 |
| 29 | Female | 71-75 | 76.67 | 16.8 | 27.9 | 121.2 | 0.44 | 273.2 | 17.8 | 24.9 | 70.3 | 0.29 | 241.3 | 8.54 |
| 30 | Male | 71-75 | 75.00 | 12.8 | 19.1 | 42.9 | 0.36 | 117.7 | 20.1 | 28.7 | 89.5 | 0.69 | 129.5 | 6.34 |
| 31 | Female | 76-80 | 73.33 | 15.2 | 24.9 | 94.2 | 0.57 | 163.9 | 12.6 | 20.6 | 78.3 | 0.49 | 160.5 | 6.96 |
| 32 | Male | 61-65 | 63.33 | 19.4 | 26.7 | 112.2 | 0.77 | 144.9 | 10.6 | 18.0 | 90.2 | 0.58 | 155.4 | 6.03 |
| 33 | Female | 71-75 | 65.00 | 9.1 | 16.7 | 55.0 | 0.36 | 153.6 | NR | NR | NR | NR | NR | 6.96 |
| 34 | Male | 66-70 | 73.33 | NR | NR | NR | NR | NR | NR | NR | NR | NR | NR | 6.55 |

## Notes

### Competing Interest Statement

The authors have declared no competing interest.

### Clinical Protocols

https://bmjopen.bmj.com/content/10/9/e039601.long

### Author Declarations

Ethics committee of the University Hospital of Antwerp, Belgium gave ethical approval for this work. (EC number B300201938949)

### Summary of Updates

Section on the role of saccular function was updated to take the effect of age into account; Figure 1 and Figure 2 are provided.

